# Accuracy of the Veterans Health Administration COVID-19 (VACO) Index for predicting short-term mortality among 1,307 Yale New Haven Hospital inpatients and 427,224 Medicare patients

**DOI:** 10.1101/2021.01.01.20249069

**Authors:** Joseph T. King, James S. Yoon, Zachary M. Bredl, Joseph P. Habboushe, Graham A. Walker, Christopher T. Rentsch, Janet P. Tate, Nitu M. Kashyap, Richard C. Hintz, Aneesh P. Chopra, Amy C. Justice

## Abstract

**Background:** The Veterans Health Administration COVID-19 (VACO) Index incorporates age, sex, and pre-existing comorbidity diagnoses readily available in the electronic health record (EHR) to predict 30-day all-cause mortality in both inpatients and outpatients infected with SARS-CoV-2. We examined the performance of the Index using data from Yale New Haven Hospital (YNHH) and national Medicare data overall, over time, and within important patient subgroups.

**Methods and findings:** With measures and weights previously derived and validated in a national Veterans Healthcare Administration (VA) sample, we evaluated the accuracy of the VACO Index for estimating inpatient (YNHH) and both inpatient and outpatient mortality (Medicare) using area under the receiver operating characteristic curve (AUC) and comparisons of predicted versus observed mortality by decile (calibration plots). The VACO Index demonstrated similar discrimination and calibration in both settings, over time, and among important patient subgroups including women, Blacks, Hispanics, Asians, and Native Americans. In sensitivity analyses, we allowed component variables to be re-weighted in the validation datasets and found that weights were largely consistent with those determined in VA data. Supplementing the VACO Index with body mass index and race/ethnicity had no effect on discrimination.

**Conclusion:** Among COVID-19 positive individuals, the VACO Index accurately estimates risk of short-term mortality among a wide variety of patients. While it modestly over-estimates risk in recent intervals, the Index consistently identifies those at greatest relative risk. The VACO Index could identify individuals who should continue practicing social distancing, help determine who should be prioritized for vaccination, and among outpatients who test positive for SARS-CoV-2, indicate who should receive greater clinical attention or monoclonal antibodies.

## Introduction

The Centers for Disease Control and Prevention (CDC) Advisory Committee on Immunization Practices (ACIP) has prioritized those 75 years and older and those 65-74 years of age with “high risk” medical conditions for novel coronavirus (COVID-19) vaccination [1]. However, these criteria identify a large proportion of the US adult population. A more tailored risk estimation incorporating specific age and pre-existing conditions could facilitate multiple objectives including: 1) motivating high risk individuals and their contacts to practice social distancing until vaccinated, 2) identification of individuals testing positive at drive up sites who require clinical examination and possibly laboratory evaluation, 3) prioritization for early outpatient monoclonal antibodies, and 4) prioritization for vaccination.

Hundreds of publications have identified risk factors associated with adverse COVID-19 outcomes, but just a small subset have developed and prospectively validated predictive models in large samples. Most of these require data available only at acute presentation (symptoms, exam findings, diagnostic imaging, or laboratory data). We find four published, internally validated, predictive models for COVID-19 mortality based exclusively on pre-existing conditions that might address the applications discussed above. The United Kingdom QCOVID model is based on a national sample of 8.25 million individuals, but concatenates testing positive, a rapidly changing risk largely dependent on ever changing local prevalence, and mortality given infection, a more biological and stable construct [2]. The study from Mexico represents 90,000 COVID-19 patients nationwide but achieves limited mortality discrimination when restricted to pre-existing conditions [3]. The Canadian model is limited to the providence of Ontario and excluded 84% of COVID-19 cases due to missing data [4].

The Veterans Health Administration COVID-19 (VACO) Index for short term mortality is based on age, sex, and comorbid diagnoses [5]. It was developed based on 3,681 SARS-CoV-2 positive patient records from the Veterans Healthcare Administration (VA) national electronic health record (EHR) and prospectively validated in 9,642 veterans split into two temporally distinct samples. Within VA, the VACO Index demonstrated good discrimination across time intervals and among men and women, White, Black, and Hispanic patients, and those living in different geographic regions. The Index is available as a web application at MDCalc.com and as a mobile application. In the first 5 weeks after the VACO Index was posted on MDCalc.com it was accessed 8,350 times. An EHR decision support tool within Epic⍰(Epic Systems Corp., Madison, WI), which auto-populates Index data elements, is currently in development.

Some have questioned the generalizability of VA data for understanding risk of mortality from COVID-19. Veterans in care are predominantly male, older, and have a higher prevalence of chronic health conditions and risk behaviors than the general US population [6-8]. To address these concerns, we compare the accuracy (discrimination and calibration) of VACO Index short term mortality estimates for COVID-19 patients in 1) our original VA sample of 13,323 inpatients and outpatients, 2) 1,307 patients admitted to Yale New Haven Hospital (YNHH, a tertiary care academic medical center), and 3) 441,854 Medicare recipients (age 65 and older). We consider the accuracy of the Index overall and within important subgroups defined by calendar time, age, sex, race/ethnicity, and geographic region. We also consider whether different weighting of the index components or adding race/ethnicity or body mass index (BMI) improves the accuracy of the VACO Index in these cohorts.

## Methods

### VACO Index components

The VACO Index, a 30-day all-cause mortality prediction model developed in VA nationwide data, utilizes demographic and pre-existing condition data available in EHR or medical administrative data [5]. The Index includes age, sex, multimorbidity quantified with the Charlson Comorbidity Index derived from International Classification of Diseases, 10th edition (ICD-10) diagnosis codes [9, 10] (S1. Table), and myocardial infarction or peripheral vascular disease – neither race nor any other individual comorbid diagnosis provided additional discriminatory function (S1. File). The VACO Index had an area under the receiver operating characteristic curve (AUC) of 0.79 in development, 0.81 in early validation, 0.84 in later validation, and validated well in sex, race/ethnicity, and regional subgroups.

### Data source and participants

#### VA data

The VACO Index was developed and validated on a nationwide sample of Veterans who were alive as of January 1, 2020 and active in care between January 1, 2018 and December 31, 2019. The Index included 13,323 individuals testing positive for SARS-CoV-2 in the inpatient or outpatient setting between March 2 and July 19, 2020 and followed them for 30 days. Data were split into a development cohort (positive test between March 2 and April 15, 2020), an early validation cohort (positive test between April 16 and May 18, 2020), and a later validation cohort (positive test between May 19 and July 19, 2020). Deaths were determined using inpatient records and the VA death registry to capture deaths occurring outside hospitalization.

#### Yale New Haven Hospital data

We extracted data from the YNNH Epic⍰EHR on 1,307 patients testing positive for SARS-CoV-2 between March 2 and July 19, 2020 who were admitted to the hospital. Patients were eligible for inclusion if they tested positive within 14 days before an admission, on the day of admission, or while hospitalized. Similar to the VACO Index methodology, we used the first SARS-CoV-2 positive test date for individuals with more than one positive test and applied the same rules for determining study baseline (see above).

Data were split into “early” and “later” cohorts temporally synchronized with the VACO Index development cohort (positive test between March 2 and April 15, 2020), and combined Index validation early and later cohorts (positive test between April 16 and July 19, 2020) cohorts, respectively. Deaths were determined by index hospitalization discharge status and post-discharge deaths captured by the health care system EHR. We used the same 30-day follow-up window for determining death as used in the VACO Index, but the YNHH data was restricted to inpatient deaths during the YNHH index and subsequent admissions, and any outpatient deaths recorded in the YNHH EHR, so we did not have complete 30-day mortality capture as in the VA data.

We grouped the YNHH data into identical age, race, and sex strata as in the VA dataset, and used the same ICD-10 diagnosis codes and algorithms to define, calculate, and group Charlson Comorbidity Index comorbidities. We also considered the most recent BMI recorded between 730 and 15 days before baseline.

#### Medicare data

We used Medicare Fee for Service data to identify 427,224 patients nationwide age 65 or older with an inpatient or outpatient COVID-19 diagnosis between March 2 and July 19, 2020, using ICD-10 diagnosis code of B97.29 on pre-April data, and U07.1 stating April 1, 2020. If an individual had a COVID-19 diagnosis code on more than one date, we only included the first date. As with the YNHH data, Medicare data were split into “early” and “late” cohorts temporally synchronized with the VACO Index development cohort (COVID-19 record date between March 2 and April 15, 2020), and combined Index validation early and later cohorts (COVID-19 record date between April 16 and July 19, 2020) cohorts, respectively. We used an identical 30-day follow-up window for determining death as used in the VACO Index. Deaths were ascertained by discharge status after any hospitalization, supplemented with deaths recorded in the Master Beneficiary Summary File within the Medicare database. The Centers for Medicare & Medicaid Services receives death information from several sources: Medicare claims data from the Medicare Common Working File, deaths submitted online by family members, and benefit information collected from the Railroad Retirement Board and the Social Security Administration.

We employed a similar data harmonization process to that used in the YNHH data for categorizing the Medicare data into identical age, race, sex, and strata as used in the VA data, and determining Charlson Comorbidity Index scores from ICD-10 diagnosis codes. We explored the more granular race stratification available in Medicare data (Non-Hispanic White, Non-Hispanic Black, Hispanic, Asian, North American Native, and Other).

### Statistical analyses

We used unadjusted and multivariable logistic regressions to model 30-day all-cause mortality. Multivariable models are depicted and compared using Forest plots. To account for missing BMI data in the VA and YNHH datasets (BMI was not available in the Medicare dataset), we used multiple imputation by chained equations, including all predictor and outcome variables and auxiliary variables, generating 10 imputed data sets for the development and validation cohorts. Using the *mi estimate* command suite in Stata, proportions, odds ratios (OR), and 95% confidence intervals (CI) were combined according to Rubin’s rules [11].

We report the AUC and calibration plots as assessments of the VACO Index performance and of alternative models in all three datasets. We used the VACO Index coefficients to calculate a predicted mortality for each patient in the YNHH and Medicare datasets, and then determined AUCs for the early and later testing dates, and in important subgroups: age (<65 vs 65+, except in Medicare data), sex (male vs female), race/ethnicity (Black vs non-Black), and US census region (except in YNHH data), and compared the values to those obtained in the original VACO Index development cohort. We refit models using the VACO Index variables in the VA, YNHH and Medicare datasets to understand how the model might have differed had it been developed in alternative datasets. To assess the impact of race and BMI, we also refit models supplementing VACO Index variables with race (VA, YNHH, and Medicare) and BMI (VA, YNHH – BMI is unavailable in Medicare data).

We assessed calibration plots of observed versus predicted 30-day mortality in ten (VA and Medicare data) or five (small sample size: YNNH data, VA female) strata containing equal numbers of deaths, in early and later testing cohorts and in subgroups by age, sex, race/ethnicity, and US census region. Data analyses were performed using Stata, version 15.1 (StataCorp, College Station, TX) and SAS 9.4 (SAS Institute Inc, Cary, NC). This study was conducted in compliance with the Health Insurance Portability and Accountability Act and was approved by the Institutional Review Boards of VA Connecticut Healthcare System and Yale University, both of whom granted waivers of consent. This cohort study is reported according to the Transparent Reporting of a multivariable prediction model for Individual Prognosis Or Diagnosis (TRIPOD) guidelines (S1. Checklist) [12].

## Results

### Participants

The dataset included 13,323 (VA), 1,307 (YNHH) and 427,224 (Medicare) patients for a total of 441,854 COVID-19 patients (Table 1). Compared to VA (median age 63 years) and YNHH (median age 65 years), Medicare patients (median age 78 years) were older. A higher proportion of Medicare patients were White (75%) compared to YNHH (44%) and VA (39%). YNHH had the largest proportion of Hispanic patients (21%). Women were better represented in YNHH (52%) and Medicare (58%) than in VA (9%). BMI was missing in 7% of VA patients and 22% of YNHH patients, and median BMI values were similar in VA (30.1 kg/m^2^) and YNHH (29.5 kg/m^2^) cohorts and unavailable in Medicare. Consistent with the older age of the Medicare cohort, most comorbid conditions were substantially more common, with a few exceptions: AIDS (2%) and mild liver disease (10%) were most common in VA data, and myocardial infarction was most common in YNHH (14%). Medicare patients were much less likely to have a Charlson Comorbidity Index score of zero indicating no comorbid disease (17%) versus VA (32%) or YNHH (47%).

**Table 1.**
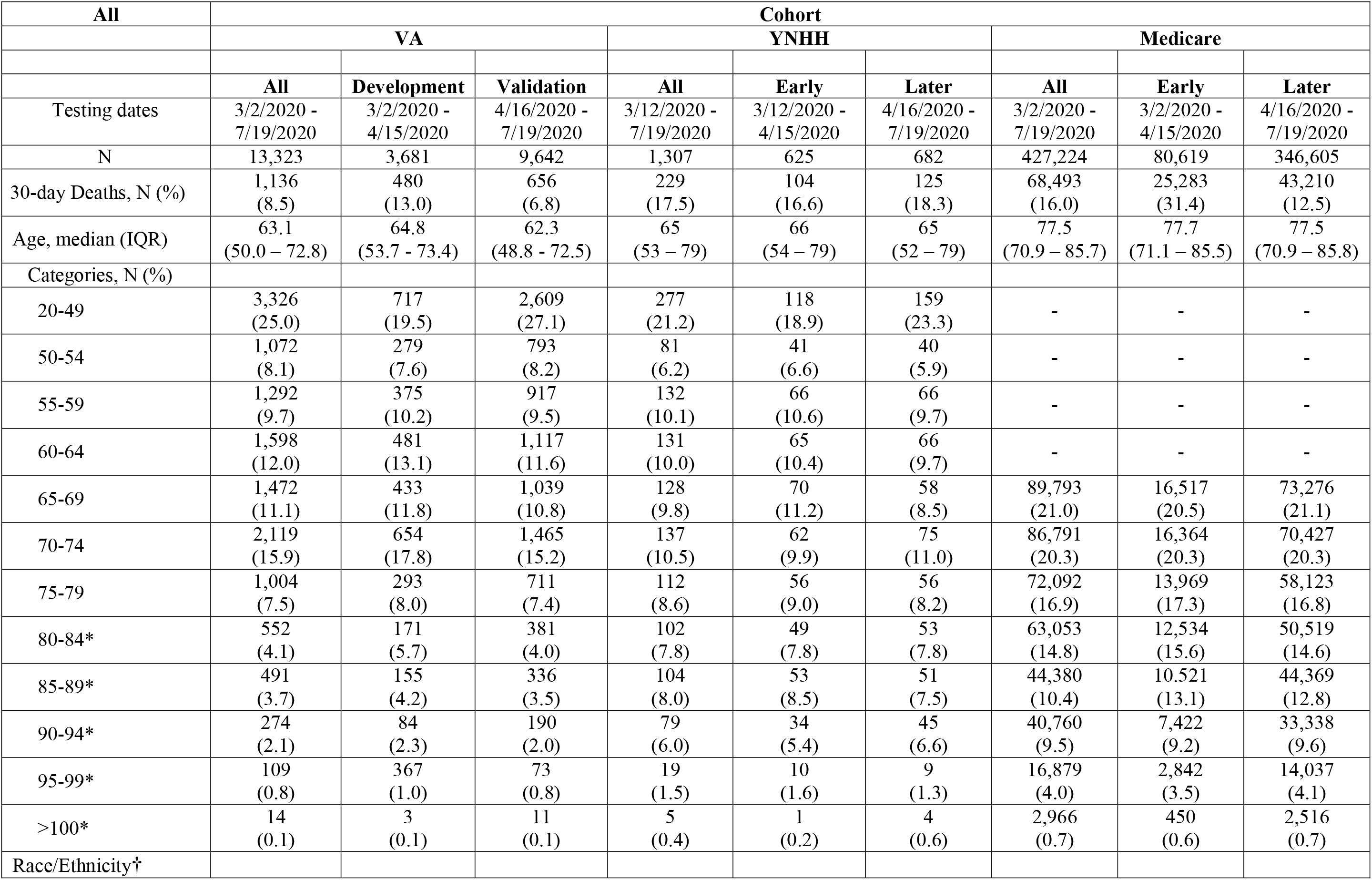
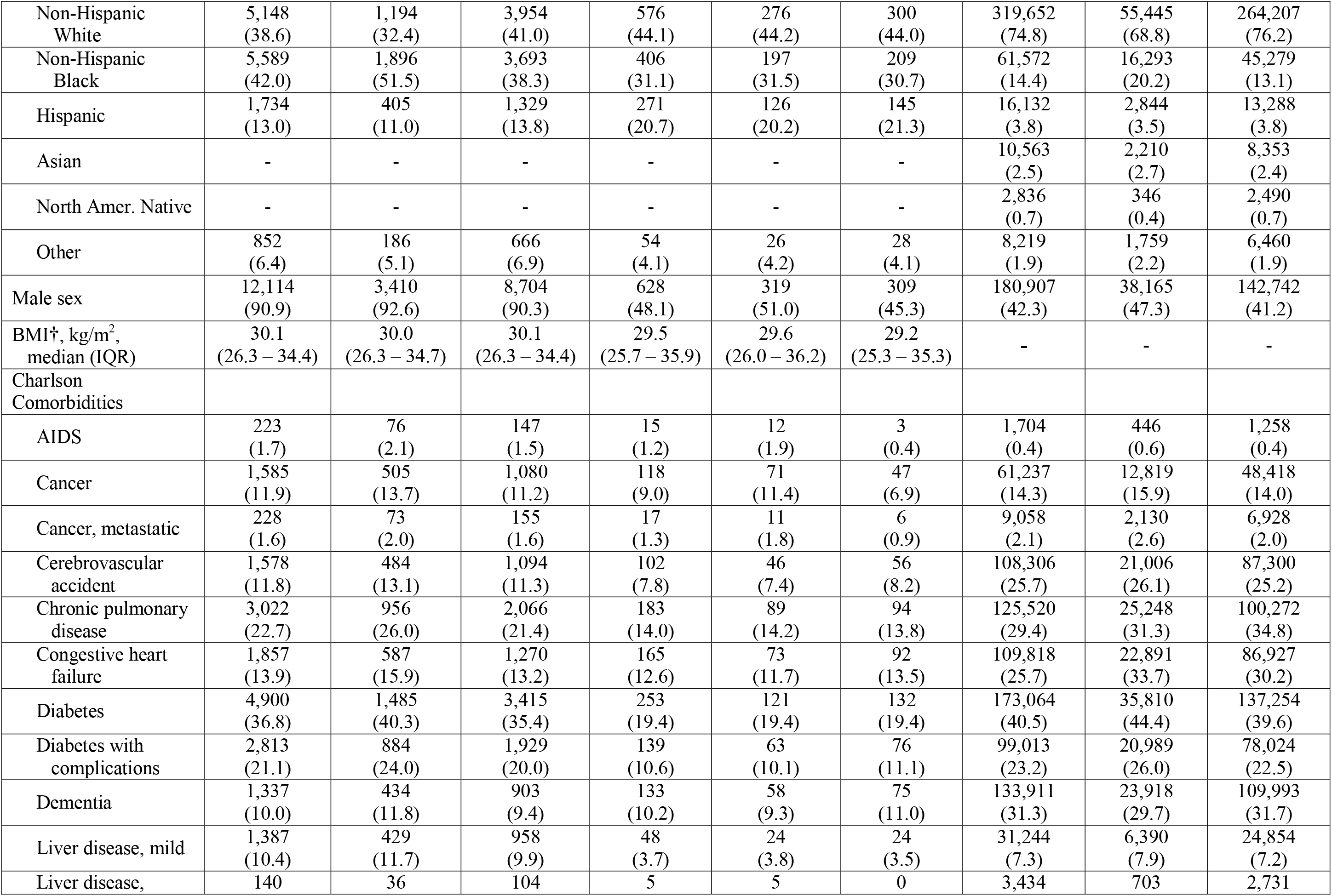
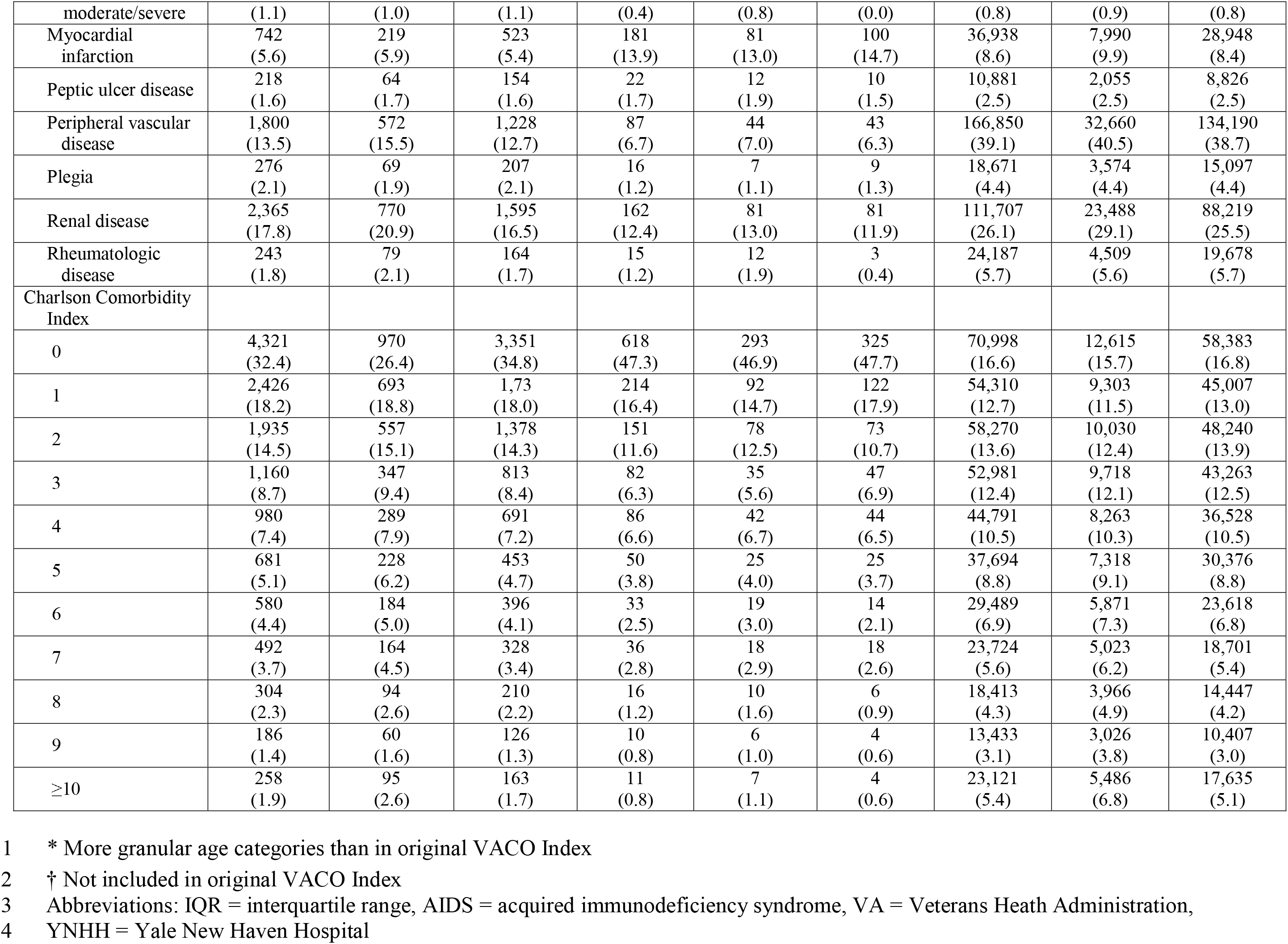
Characteristics of Patients in VA, YNHH, and Medicare Cohorts.

### Early vs later testing patients

In comparing early versus later testing period patients (Table 1), a greater proportion in the later period were in the youngest age strata of 20-49 years in both VA (early 19.5% vs later 27.1%) and YNHH (early 18.9% vs later 23.3%). A larger proportion were Hispanic in the later period in VA (early 11.0% vs later 13.8%), YNHH (early 20.2% vs later 21.2%), and Medicare (early 3.5% vs later 3.8%). Fewer in the later period were Black in VA (early 51.5% vs later 38.3%), YNHH (early 31.5% vs later 30.7%) and Medicare (early 20.2% vs later 13.1%). In Medicare, the proportion of Asians decreased (early 2.7% vs later 2.4%) and Native North Americans nearly doubled (early 0.4% vs later 0.7%). Across datasets, the proportion with specific comorbid diseases decreased and the proportion with a Charlson Comorbidity Index of 0 increased in VA (early 26.4% vs later 34.8%), in YNHH (early 46.9% vs later 47.7%), and in Medicare (early 15.7% vs later 16.8%).

### Risk factors associated with mortality

We compared unadjusted ORs estimated in each cohort among all adult COVID-19 patients in VA and YNHH (Table 2a). Additionally, since our Medicare data was limited to those 65 years of age and over, we applied this age restriction to all three cohorts in VA, YNHH and Medicare. Among all adult COVID-19 patients in VA and YNHH, VA unadjusted ORs for mortality fell within the confidence intervals of the YNHH sample for age, BMI, and most specific comorbid conditions. Increasing BMI was associated with a decreased risk of mortality in unadjusted analyses in both VA (OR: 0.97, 95% CI: 0.96 − 0.99) and YNHH (OR: 0.94, 95% CI: 0.91 − 0.97) data. Unadjusted odds of mortality associated with diabetes, diabetes with complications, and peptic ulcer disease were higher in VA data than YNHH (Table 2a). The OR for mortality associated with male sex was higher in VA (OR: 4.78, 95% CI: 3.20 – 7.14) than YNHH (OR: 1.02, 95% CI: 0.77 − 1.36) data.

**Table 2a.**
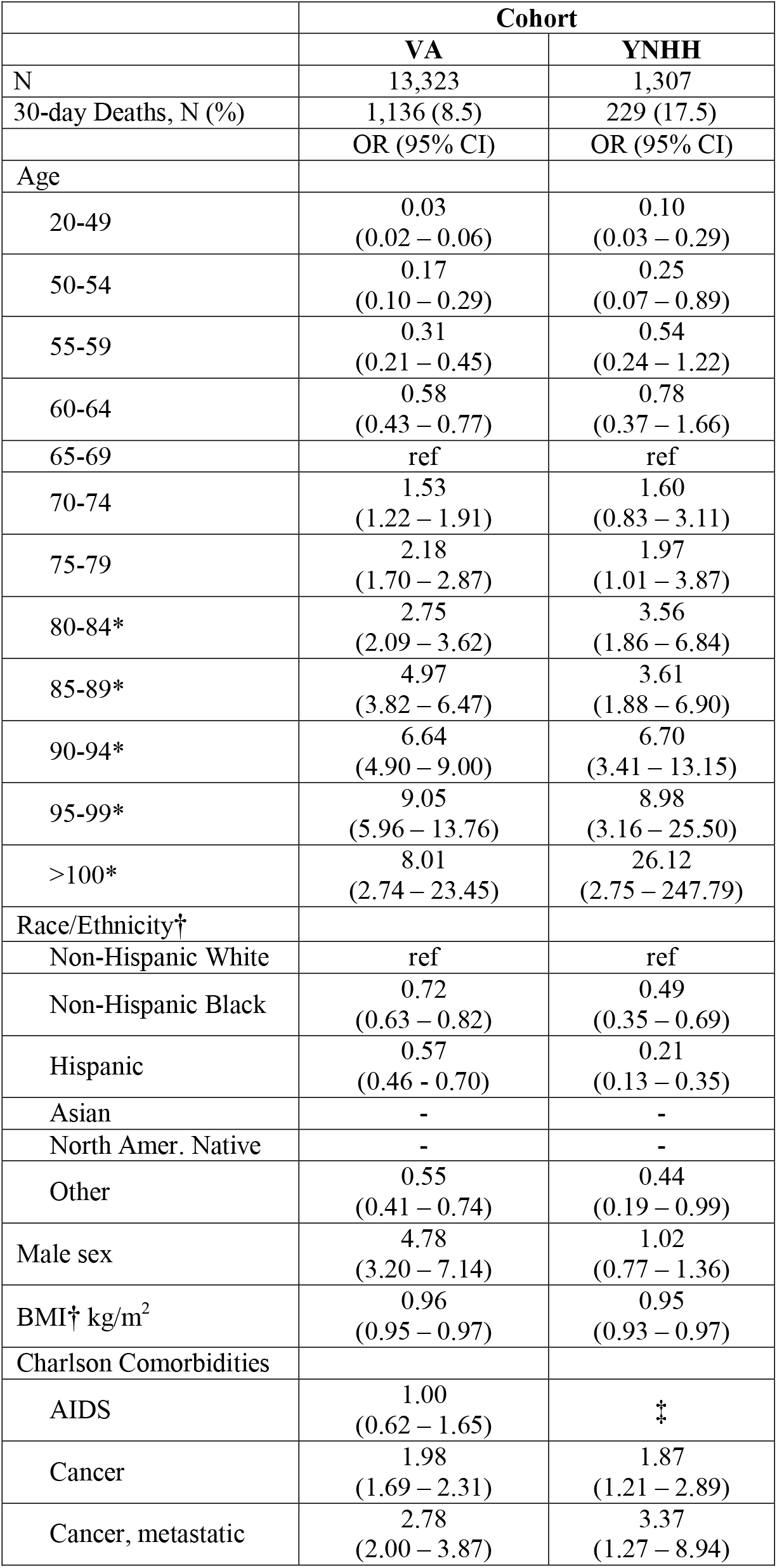
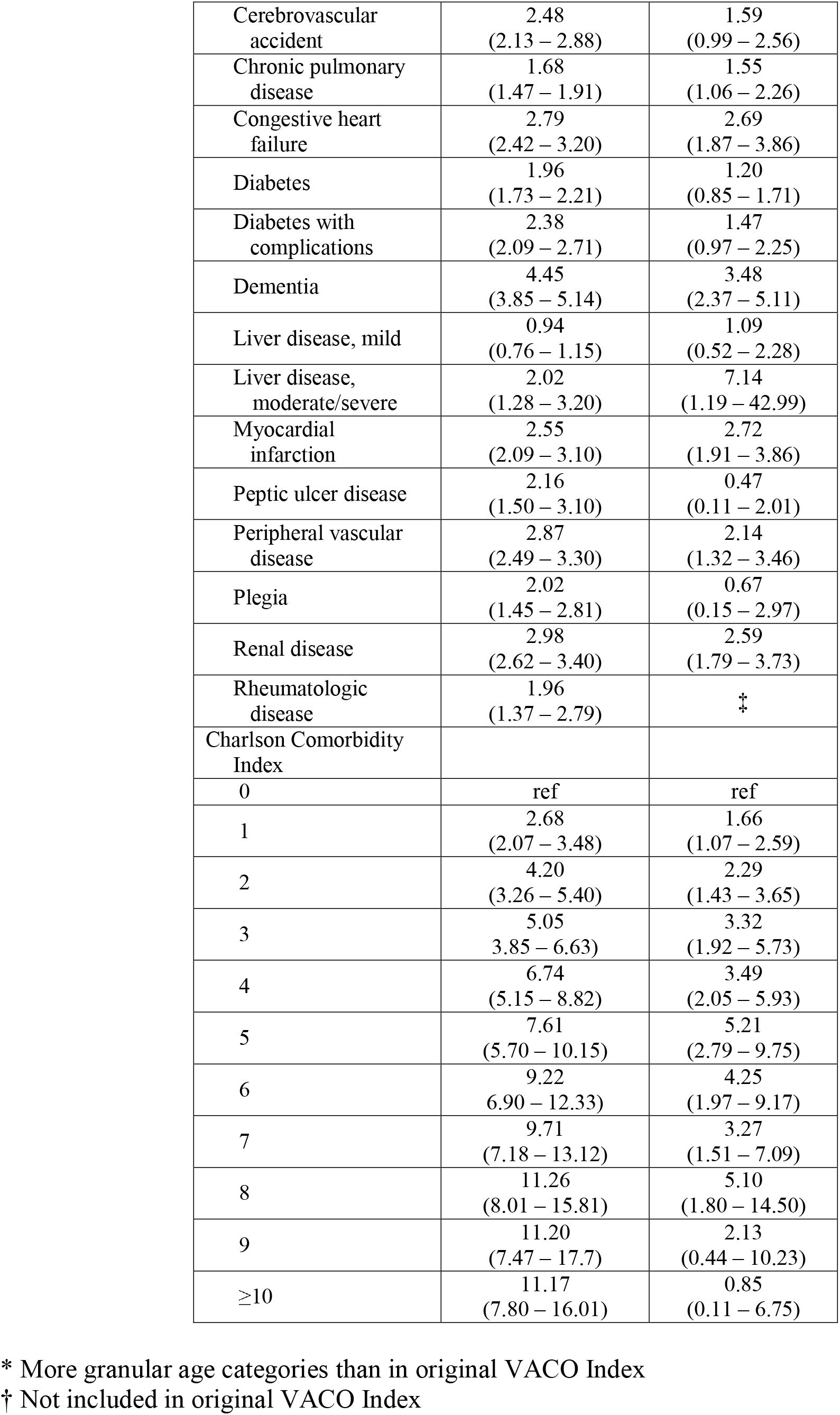
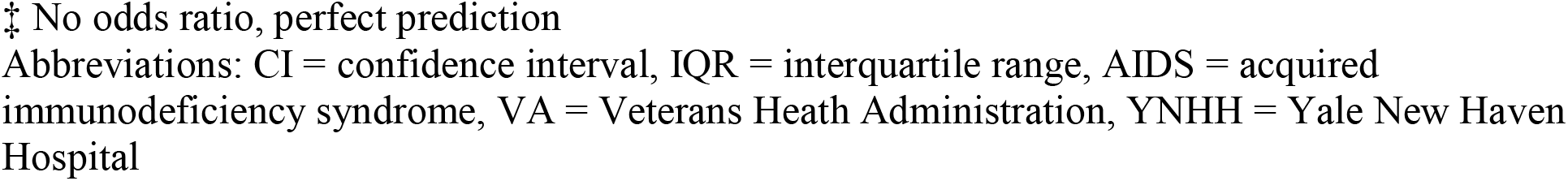
**VACO Index Components Unadjusted Associations with 30-Day Mortality in VA and YNHH Cohorts, Age 20 and Older**

**Table 2b.**
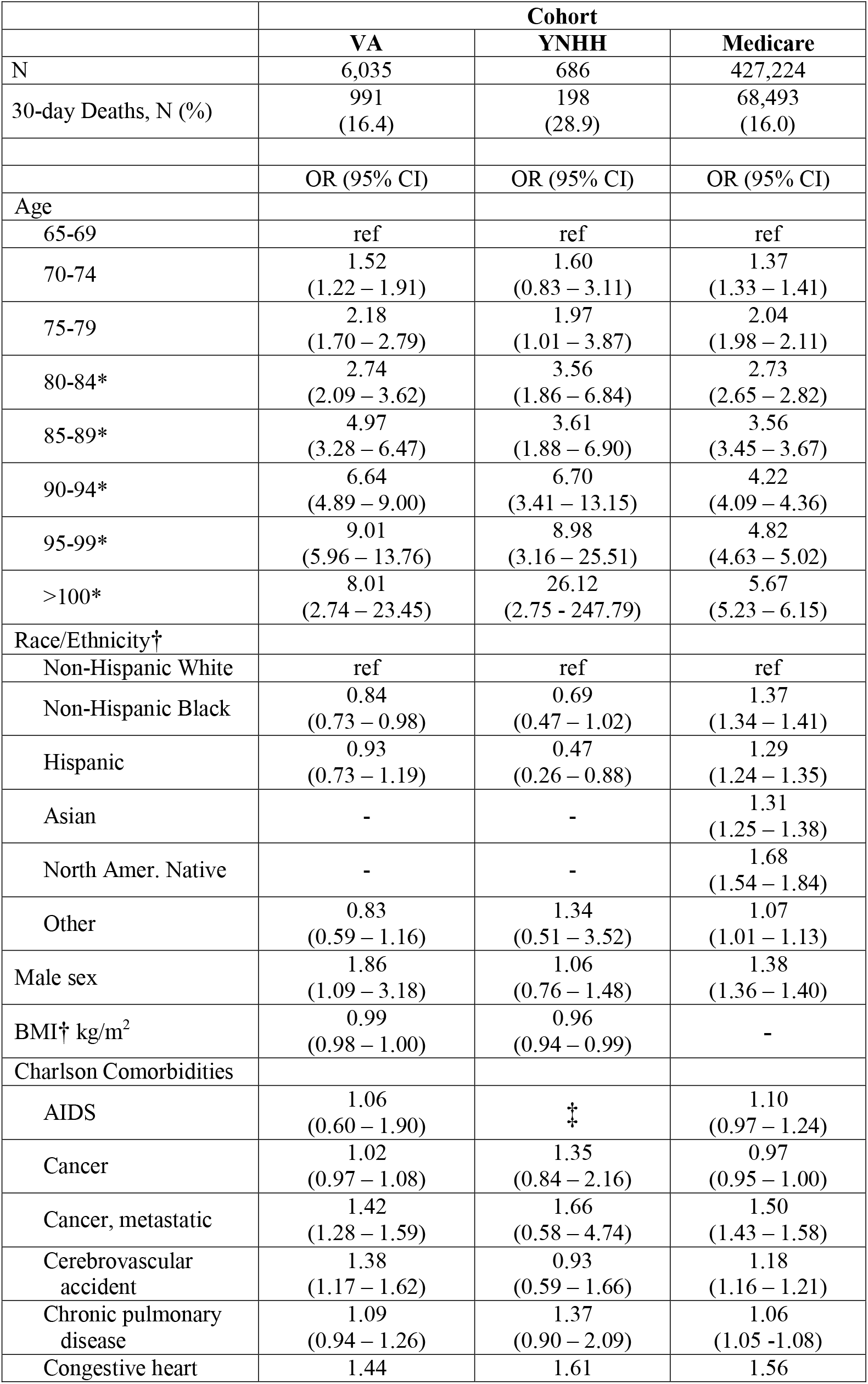
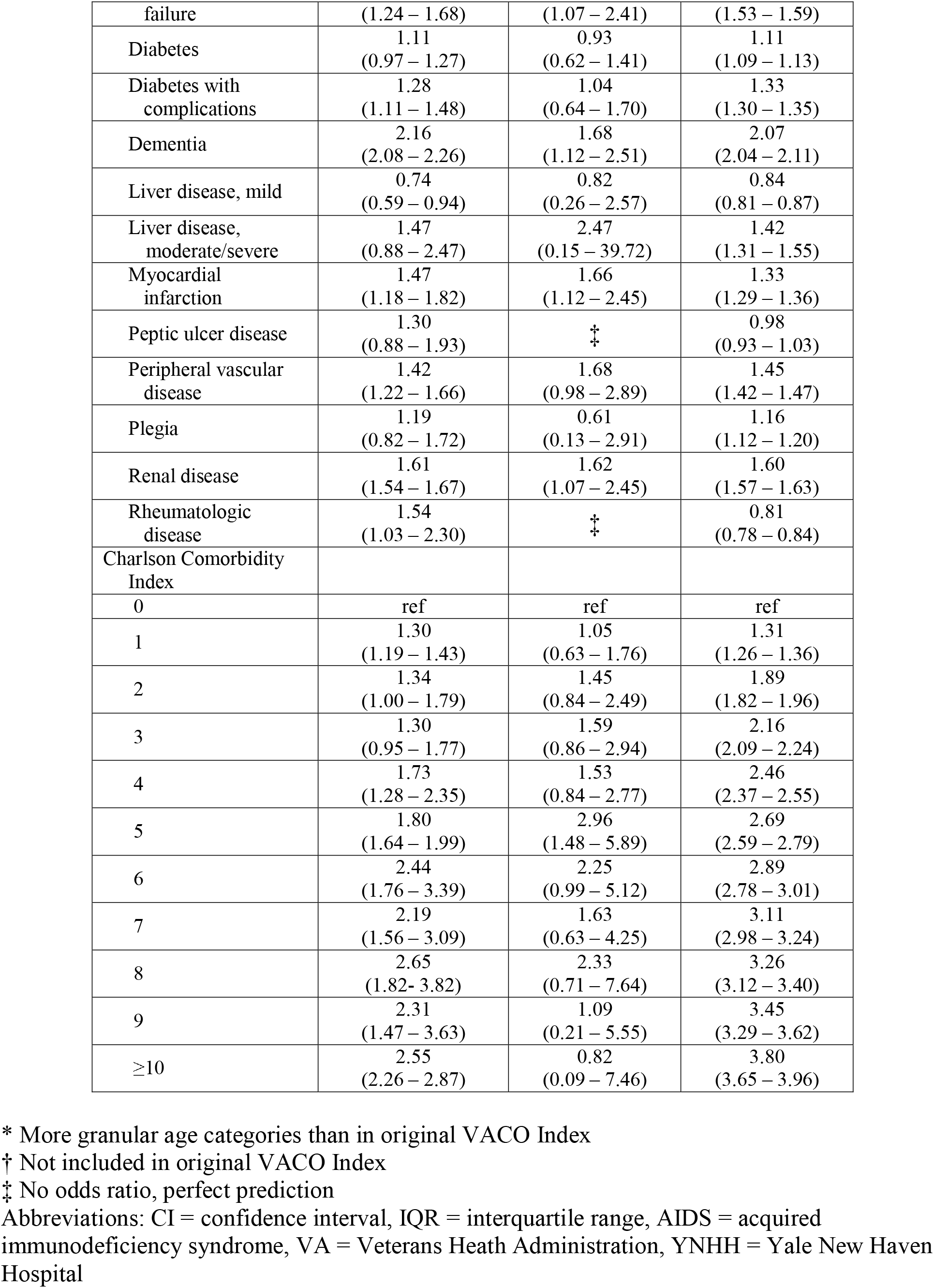
**VACO Index Components Unadjusted Associations with 30-Day Mortality in VA, YNHH, and Medicare Cohorts, Age 65 and Older**

Among those 65 years of age and over, we observed a steeper increasing gradient of ORs associated with increasing age, and a shallower increasing gradient of ORs associated with increasing Charlson Comorbidity Index scores among VA and YNHH patients than among Medicare patients (Table 2b). Black race and Hispanic ethnicity were not associated with increased mortality in VA or YNHH data, but both were associated with increased mortality in Medicare data (Black OR: 1.37, 95% CI: 1.34 − 1.41); Hispanic (OR: 1.29, 95% CI: 1.24 − 1.35). Of note, only Medicare data included a sufficient sample of Native North Americans and Asians to estimate ORs. Asians had a similar increased ORs (1.31, 95% CI: 1.25 − 1.38) and Native North Americans had a higher ORs for mortality (1.68, 95% CI: 1.54 − 1.84). In contrast to the overall adult sample of VA and YNHH, among those 65 years and older, ORs for male sex and mortality were consistent across cohorts. Similarly, ORs for mortality estimated for specific comorbid conditions were largely consistent across cohorts with the exception of dementia which was weighted more heavily in VA (OR: 2.16, 95% CI: 2.08 − 2.26) than in Medicare (OR: 2.08, 95% CI: 2.04 − 2.11) and rheumatologic disease which was protective in Medicare (OR: 0.81, 95% CI: 0.78 − 0.84) and associated with increased risk of mortality in VA (OR: 1.54, 95% CI: 1.03 − 2.30).

### Performance in the YNHH cohort

The VACO Index demonstrated good discrimination in YNHH data overall (AUC: 0.80, 95% CI: 0.77 − 0.83), consistent with that seen in VA overall (AUC: 0.82, 95% CI: 0.81 − 0.83) (Table 3a). Discrimination in YNHH early and later periods were similar to the VA development period (Table 3a). When AUCs were estimated for subgroups, the VACO Index demonstrated similar discrimination in YNHH data as in VA data among those <65 years of age, those 65 and over, men, Non-Hispanic Black patients, Hispanic patients and Other race/ethnicity patients. The discrimination among Non-Hispanic White patients at YNHH was not as strong (AUC: 0.72, 95% CI: 0.67 − 0.77) as in VA (AUC: 0.82, 95% CI: 0.81 − 0.84). While good, discrimination among women in YNHH (AUC: 0.81, 95% CI: 0.77 − 0.85) was not as strong as among women in VA (AUC: 0.88, 95% CI: 0.81 − 0.95).

**Table 3a.** **Validation of VACO Index 30-day COVID-19 Mortality Estimates in Subgroups of VA and YNHH Cohorts**

|  | **VA** | **YNHH** |
| --- | --- | --- |
|  | AUC<br>(95% CI) | AUC<br>(95% CI) |
| N | 13,323 | 427,224 |
| 30-day Deaths, N (%) | 1,136<br>(8.5) | 68,493<br>(16.0) |
| Overall | 0.82<br>(0.81 – 0.83) | 0.80<br>(0.77 – 0.83) |
| Subgroups |  |  |
| Testing Date |  |  |
| Early, 3/2 – 4/15 | 0.79<br>(0.77 – 0.81) | 0.82<br>(0.78 – 0.86) |
| Later, 4/16 – 7/19 | 0.84<br>(0.82 – 0.85) | 0.78<br>(0.74 – 0.82) |
| Age |  |  |
| <65 | 0.80<br>(0.77 – 0.84) | 0.77<br>(0.69 – 0.85) |
| 65+ | 0.69<br>(0.67 – 0.70) | 0.68<br>(0.64 – 0.73) |
| Sex |  |  |
| Male | 0.81<br>(0.80 – 0.82) | 0.79<br>(0.75 – 0.83) |
| Female | 0.88<br>(0.81 – 0.95) | 0.81<br>(0.77 – 0.85) |
| Race/Ethnicity |  |  |
| Non-Hispanic White | 0.82<br>(0.81 – 0.84) | 0.72<br>(0.67 – 0.77) |
| Non-Hispanic Black | 0.80<br>(0.78 – 0.81) | 0.81<br>(0.75 – 0.86) |
| Hispanic | 0.85<br>(0.82 – 0.88) | 0.87<br>(0.82 – 0.95) |
| Other | 0.87<br>(0.83 – 0.91) | 0.94<br>(0.87 – 1.00) |

Abbreviations: AUC = Area under receiver operating characteristic curve, CI = confidence interval, YNHH = Yale New Haven Hospital

**Table 3b.**
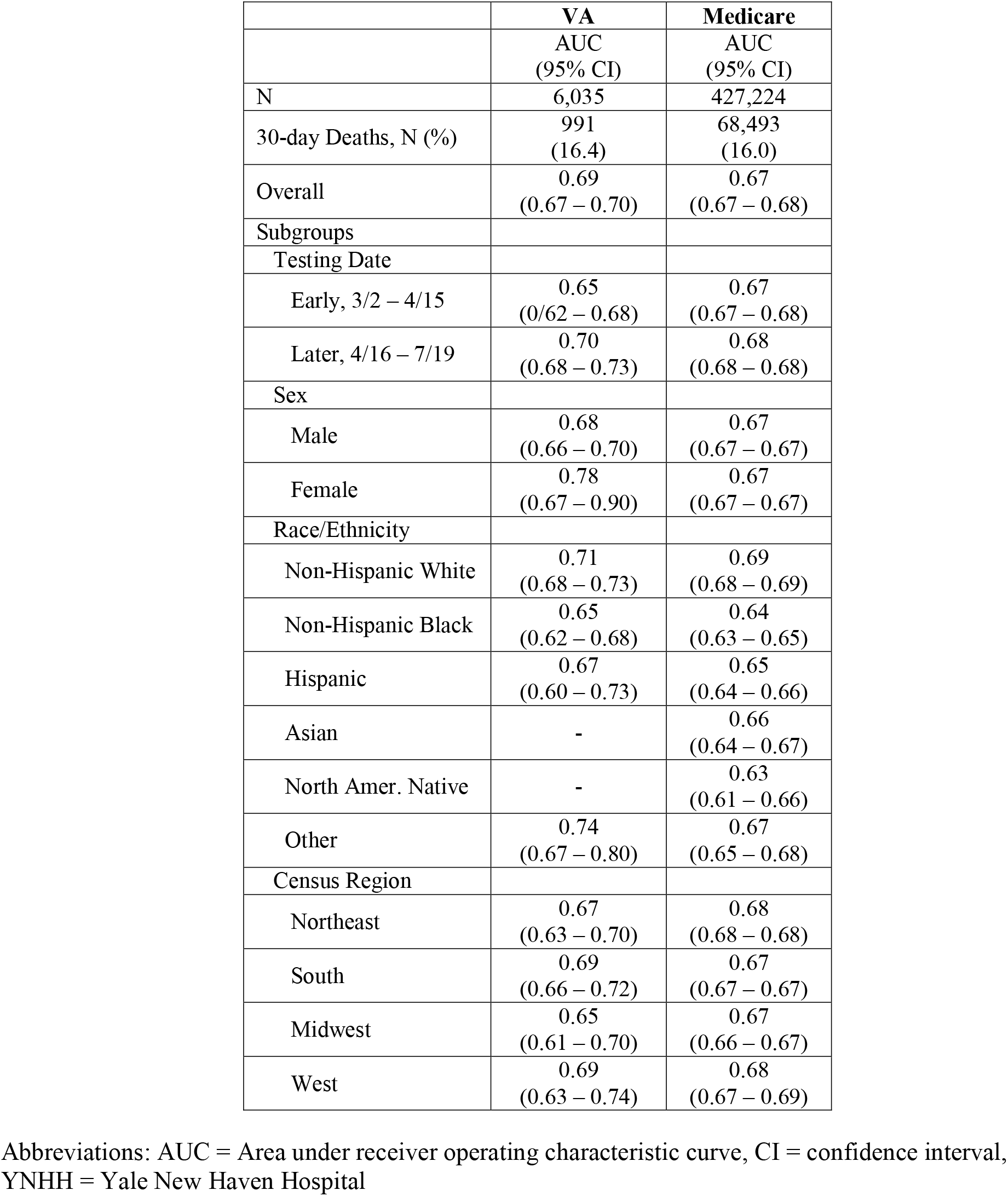
**Validation of VACO Index 30-day COVID-19 Mortality Estimates in Subgroups of VA and Medicare Cohorts, Age 65 and Over**

|  | **VA** | **Medicare** |
| --- | --- | --- |
|  | AUC<br>(95% CI) | AUC<br>(95% CI) |
| N | 6,035 | 427,224 |
| 30-day Deaths, N (%) | 991<br>(16.4) | 68,493<br>(16.0) |
| Overall | 0.69<br>(0.67 – 0.70) | 0.67<br>(0.67 – 0.68) |
| Subgroups |  |  |
| Testing Date |  |  |
| Early, 3/2 – 4/15 | 0.65<br>(0.62 – 0.68) | 0.67<br>(0.67 – 0.68) |
| Later, 4/16 – 7/19 | 0.70<br>(0.68 – 0.73) | 0.68<br>(0.68 – 0.68) |
| Sex |  |  |
| Male | 0.68<br>(0.66 – 0.70) | 0.67<br>(0.67 – 0.67) |
| Female | 0.78<br>(0.67 – 0.90) | 0.67<br>(0.67 – 0.67) |
| Race/Ethnicity |  |  |
| Non-Hispanic White | 0.71<br>(0.68 – 0.73) | 0.69<br>(0.68 – 0.69) |
| Non-Hispanic Black | 0.65<br>(0.62 – 0.68) | 0.64<br>(0.63 – 0.65) |
| Hispanic | 0.67<br>(0.60 – 0.73) | 0.65<br>(0.64 – 0.66) |
| Asian | - | 0.66<br>(0.64 – 0.67) |
| North Amer. Native | - | 0.63<br>(0.61 – 0.66) |
| Other | 0.74<br>(0.67 – 0.80) | 0.67<br>(0.65 – 0.68) |
| Census Region |  |  |
| Northeast | 0.67<br>(0.63 – 0.70) | 0.68<br>(0.68 – 0.68) |
| South | 0.69<br>(0.66 – 0.72) | 0.67<br>(0.67 – 0.67) |
| Midwest | 0.65<br>(0.61 – 0.70) | 0.67<br>(0.66 – 0.67) |
| West | 0.69<br>(0.63 – 0.74) | 0.68<br>(0.67 – 0.69) |

Abbreviations: AUC = Area under receiver operating characteristic curve, CI = confidence interval, YNHH = Yale New Haven Hospital

### Performance in the Medicare cohort

Restricting all cohorts to the age range included in Medicare (65 years or older), the VACO Index demonstrated consistent AUCs in VA and Medicare data (Table 3b). In all cases, the AUCs estimated within Medicare data fell within the 95% CI for the VA AUCs overall and by testing date, gender, b race/ethnicity and census region.

When comparing the age-restricted VA and Medicare calibration plots, there was evidence of modest over prediction in both samples (Figures 2a and b). Of note, the Index slightly under-estimated risk in the early interval of Medicare data but was better calibrated among Black, Hispanic, Asian, and North American Native than among White patients.

**Figure 1a.**
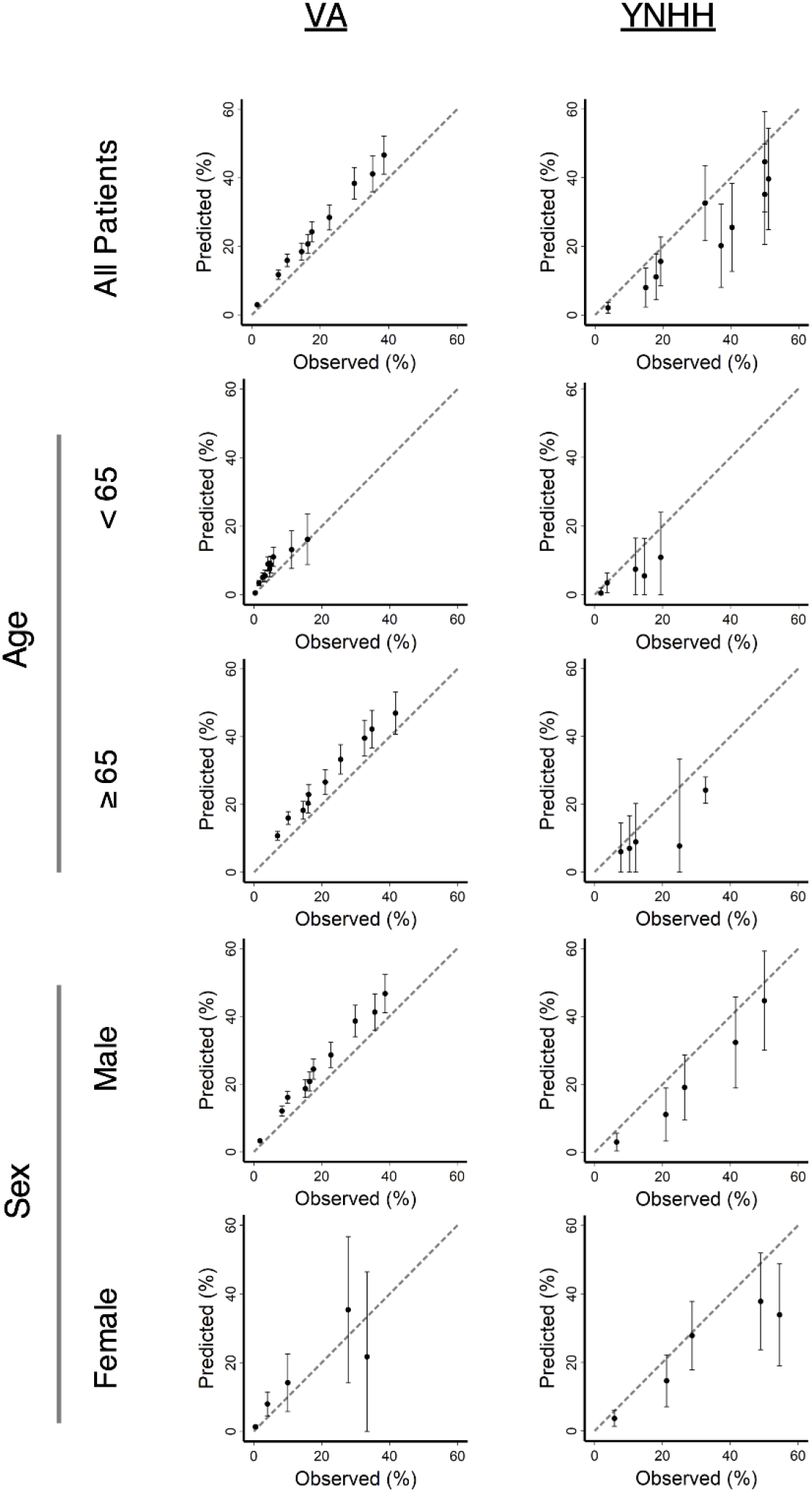
Calibration plots comparing VACO Index predicted and observed 30-day mortality in Veterans Health Administration (VA) and Yale New Haven Health (YNHH) data, overall and in age and sex subgroups. The dashed diagonal line represents perfect prediction. Values above the dashed line indicate overprediction of mortality by the VACO Index, and values below the line represent underprediction. Error bars depict 95% confidence intervals of mortality predictions. Small VA female overall YNHH data sample size limited subgroup plots to five strata and produced wide confidence intervals.

**Figure 1b.**
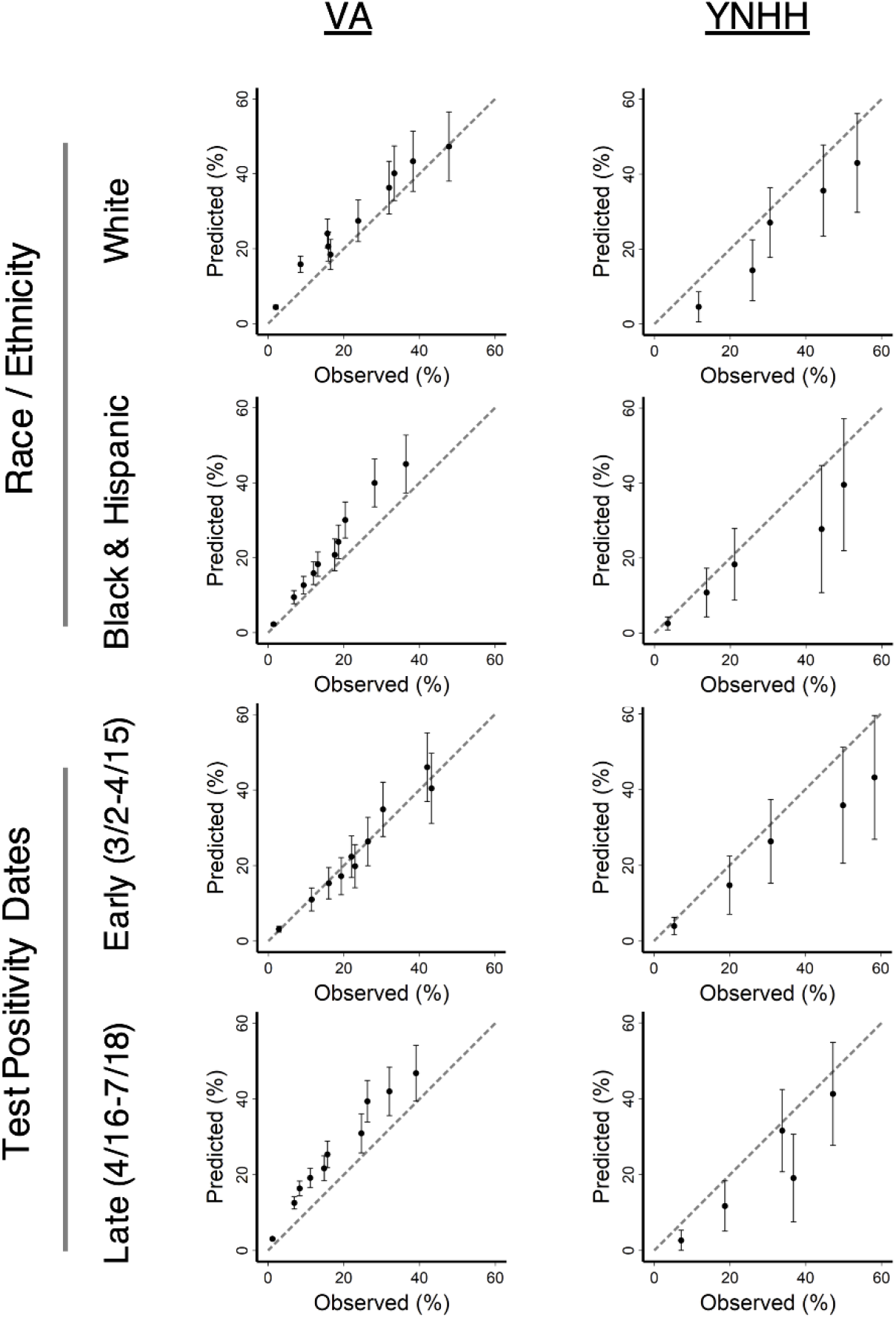
Calibration plots comparing VACO Index predicted and observed 30-day mortality in Veterans Health Administration (VA) and Yale New Haven Health (YNHH) data in Non-Hispanic White vs Non-Hispanic Black and Hispanic subgroups, and in early (March 2 to April 15, 2020) vs later (April 16 – July 19, 2020) testing date subgroups. The dashed diagonal line represents perfect prediction. Values above the dashed line indicate overprediction of mortality by the VACO Index, and values below the line represent underprediction. Error bars depict 95% confidence intervals of mortality predictions. Small sample size of YNHH data limited subgroup plots to five strata and produced wide confidence intervals.

**Figure 2a.**
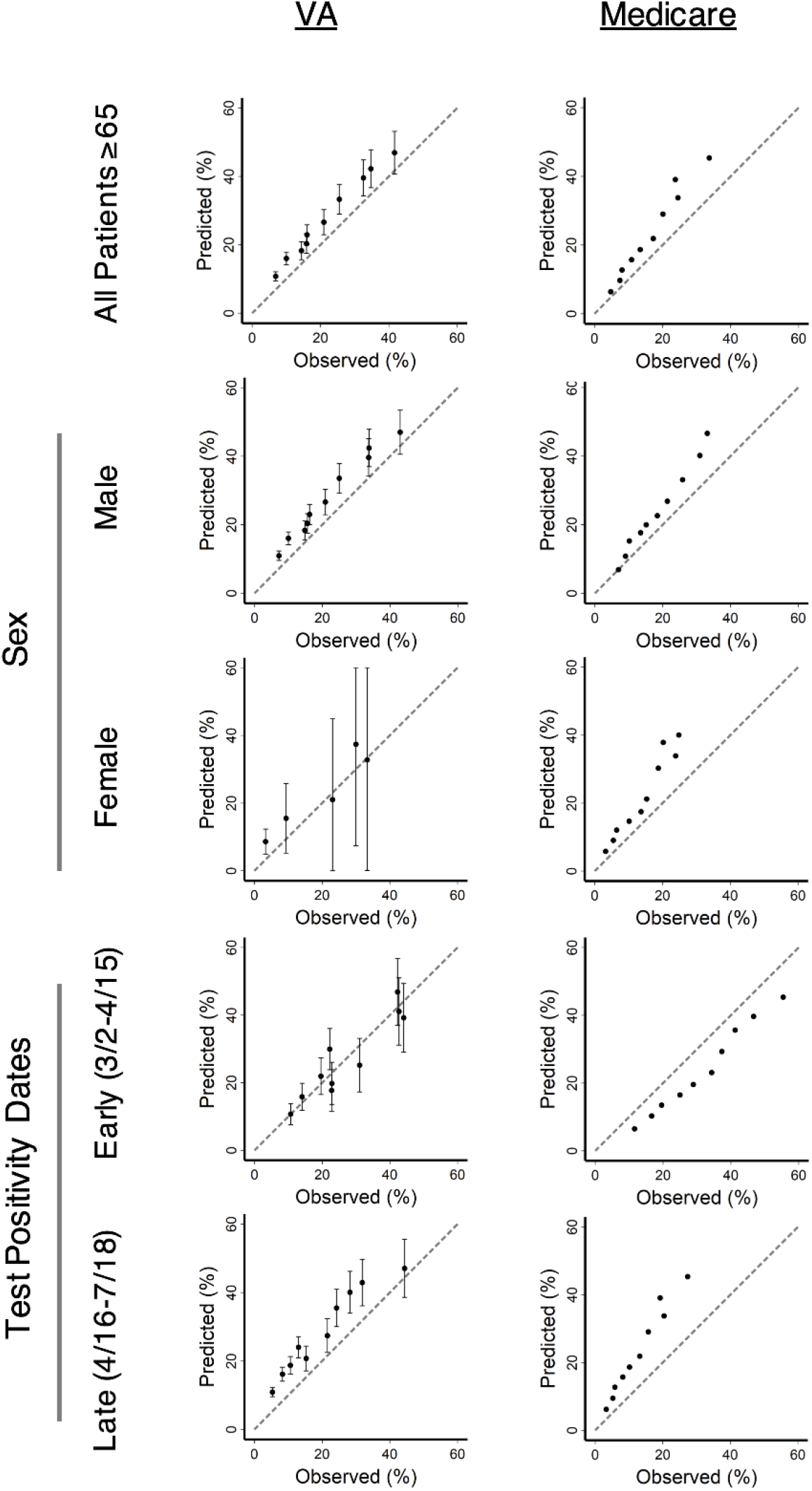
Calibration plots comparing VACO Index predicted and observed 30-day mortality in Veterans Health Administration (VA) patients age 65 and older and Medicare data, overall and in sex and early (March 2 to April 15, 2020) vs later (April 16 – July 19, 2020) testing date subgroups. The dashed diagonal line represents perfect prediction. Values above the dashed line indicate overprediction of mortality by the VACO Index, and values below the line represent underprediction. Error bars depict 95% confidence intervals of mortality predictions – large sample size of Medicare data produced tight 95% confidence intervals that are not visible at this plot scale. Small VA female sample size limited plots to five strata.

**Figure 2b.**
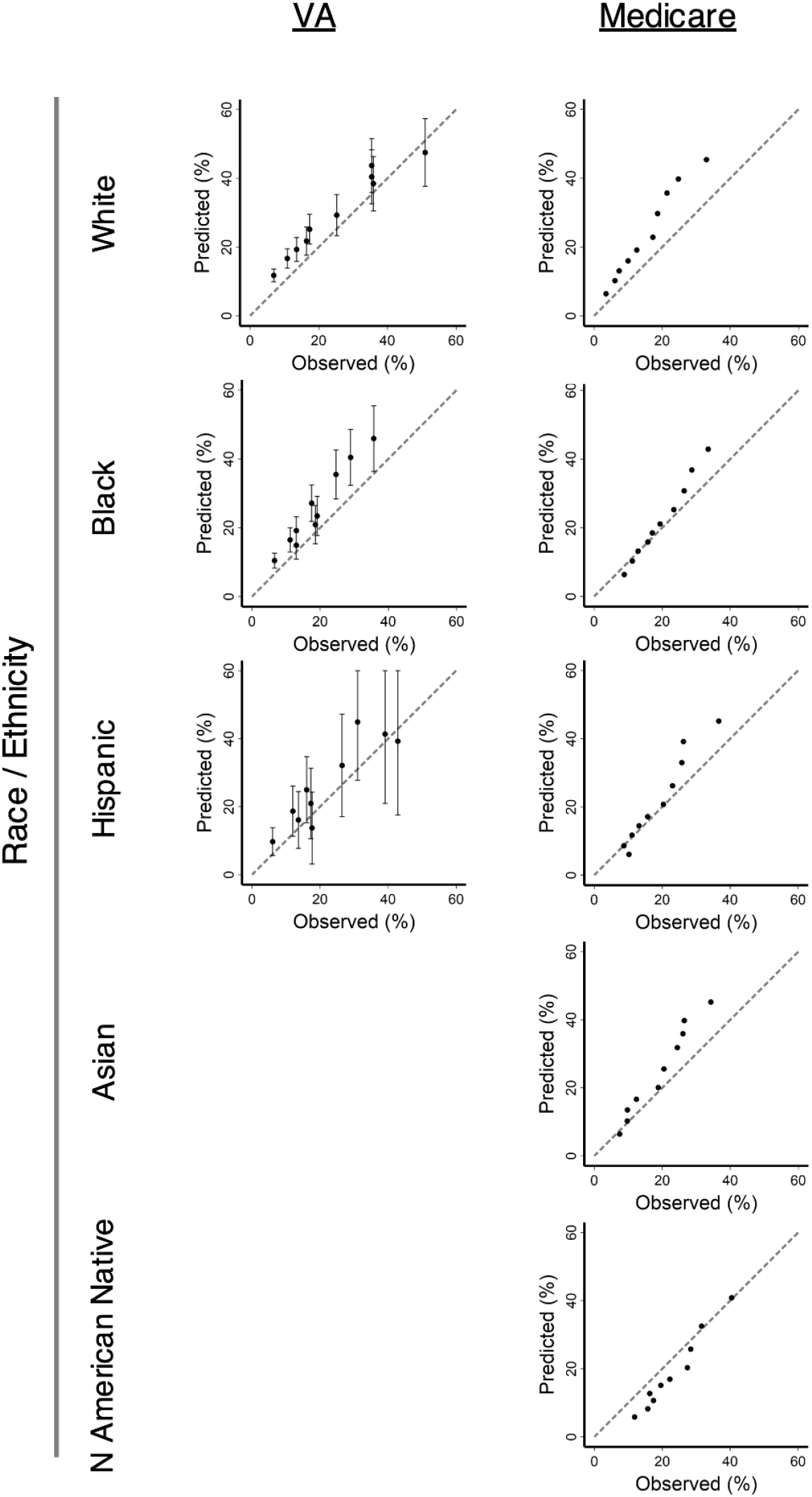
Calibration plots comparing VACO Index predicted and observed 30-day mortality in Veterans Health Administration (VA) patients age 65 and older and Medicare data in race/ethnicity subgroups: Non-Hispanic White, Non-Hispanic Black, Hispanic, Asian, and North American Natives (the latter two categories only available in Medicare data). The dashed diagonal line represents perfect prediction. Values above the dashed line indicate overprediction of mortality by the VACO Index, and values below the line represent underprediction. Error bars depict 95% confidence intervals of mortality predictions – large sample size of Medicare data produced tight 95% confidence intervals that are not visible at this plot scale.

### VACO Index supplemented with race and BMI

Compared to the original VACO Index (Table 4, model 1), the additional of race and BMI did not improve AUCs in VA, YNHH, or Medicare data (model 2). Refitting the VACO Index variables in YNHH and Medicare data improved the AUC by a single point (YNHH 0.80 to 0.81; Medicare early 0.67 to 0.68, Medicare later 0.68 to 0.69, model 3), and adding race and BMI to the refit did not improve the AUCs further (model 4). Fully adjusted model AUCs (models 5-7) fell largely within the confidence intervals estimated in the earlier models. Forest plots of ORs from the VACO Index and refitted models in VA, YNHH, and Medicare data using VACO Index variables supplemented with race and BMI showed similar ORs for age, sex, race, Charlson Comorbidity Index, MI or PVD, and BMI (Figure 3).

**Table 4.** **Impact of Adding Race and BMI to VACO Index 30-day COVID-19 Mortality Estimates in VA, YNHH, and Medicare Cohorts**

|  |  | Cohort |  |  |  |  |
| --- | --- | --- | --- | --- | --- | --- |
|  |  | VA |  | YNHH | Medicare |  |
|  |  | Development | Validation | Early and Late* | Early | Late |
| Model |  | AUC<br>(95% CI) | AUC<br>(95% CI) | AUC<br>(95% CI) | AUC<br>(95% CI) | AUC<br>(95% CI) |
| 1 | VACO Index | 0.79<br>(0.77 - 0.81) | 0.84<br>(0.82 - 0.85) | 0.80<br>(0.77 - 0.83) | 0.67<br>(0.67 - 0.68) | 0.68<br>(0.68 - 0.68) |
| 2 | VACO Index Fixed + Race + BMI | 0.79<br>(0.78 - 0.79) | 0.84<br>(0.83 - 0.84) | 0.80<br>(0.77 - 0.83) | 0.67<br>(0.67 - 0.68) | 0.67<br>(0.67 - 0.68) |
| 3 | Refit: VACO Index Variables | 0.79<br>(0.77 - 0.81) | 0.84<br>(0.83 - 0.85) | 0.81<br>(0.78 - 0.84) | 0.68<br>(0.67 - 0.68) | 0.69<br>(0.68 - 0.69) |
| 4 | Refit: VACO Index Variables + Race + BMI† | 0.79<br>(0.77 - 0.81) | 0.84<br>(0.83 - 0.85) | 0.81<br>(0.78 - 0.84) | 0.68<br>(0.68 - 0.68) | 0.68<br>(0.68 - 0.69) |
| 5 | Refit: Medicare Age, Sex, CCI 0-10, Race, BMI† | 0.79<br>(0.77 - 0.81) | 0.84<br>(0.83 - 0.85) | 0.81<br>(0.79 - 0.84) | 0.68<br>(0.68 - 0.68) | 0.68<br>(0.68 - 0.69) |
| 6 | Refit: Medicare Age, Sex, 17 CCI variables, Race, BMI† | 0.80<br>(0.78 - 0.82) | 0.85<br>(0.83 - 0.86) | 0.82<br>(0.79 - 0.84) | 0.68<br>(0.67 - 0.68) | 0.69<br>(0.68 - 0.69) |
| 7 | Refit: Medicare Age, Sex, 17 CCI variables, CCI 0-10, Race, BMI† | 0.80<br>(0.78 - 0.82) | 0.85<br>(0.83 - 0.86) | 0.82<br>(0.80 - 0.85) | 0.68<br>(0.67 - 0.68) | 0.69<br>(0.68 - 0.69) |

\* YNHH early and later cohorts combined due to small sample size
† BMI not in Medicare models
Abbreviations: AUC = Area under receiver operating characteristic curve, CI = confidence interval, VA = Veterans Health Administration, YNHH = Yale New Haven Hospital, CCI = Charlson Comorbidity Index, BMI = Body mass index

**Figure 3.**
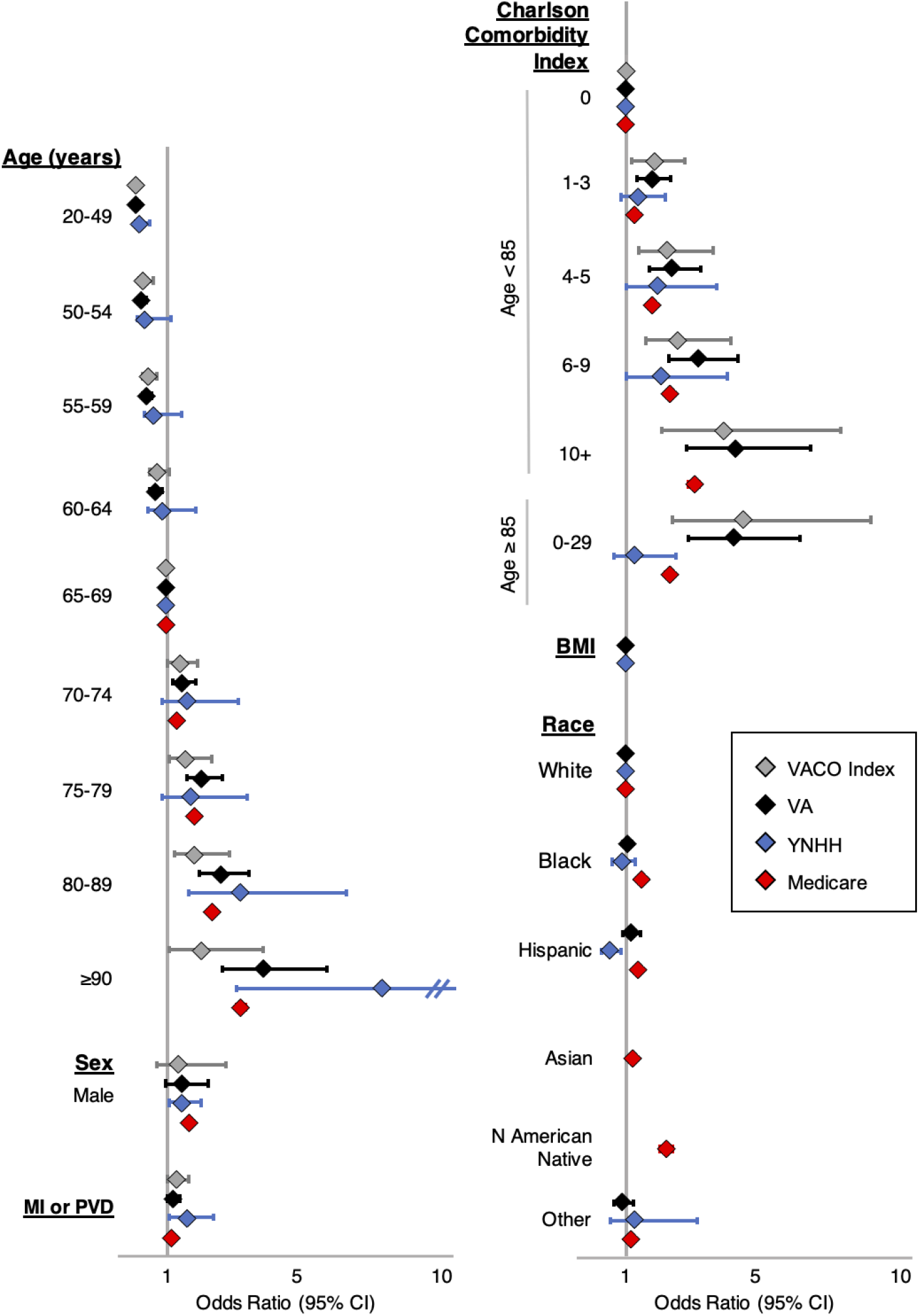
Forest plot of multivariable models of COVID-19 30-day mortality. VACO Index original model includes age, sex, Charlson Comorbidity Index and age interaction term, and myocardial infarction (MI) or peripheral vascular disease (PVD). Veterans Health Administration (VA) data model refits VACO Index variables and adds body mass index (BMI) and race (Non-Hispanic White, Non-Hispanic Black, Hispanic, Other). Yale New Haven Health (YNHH) data model refits VACO Index variables and adds BMI and additional race categories (Non-Hispanic White, Non-Hispanic Black, Hispanic, and Other). Medicare data model refits VACO Index variables, and adds BMI and race (Non-Hispanic White, Non-Hispanic Black, Hispanic, Asian North American Native, and Other). Models and odds ratios of most variables are consistent across the datasets, providing evidence of the generalizability of the VACO Index model.

## Discussion

Based on age, sex and pre-existing conditions, the VACO Index maintains good discrimination and calibration for short-term mortality among patients with COVID-19 in two demographically diverse samples outside the VA healthcare system. While unadjusted associations between specific variables and mortality vary by database, adjusted associations identified in VA data are largely consistent in YNHH and Medicare data, demonstrating that relative weights assigned variables in the VACO index are reasonably consistent with the weights that would be assigned with YNHH or Medicare data. These findings suggest that COVID-19 mortality associations seen in VA data generalize reasonably well to other United States populations. Additionally, even though mortality rates have decreased over time, the index effectively discriminates those at greater and lesser risk of mortality in both time periods within all three samples. This is true regardless of patient sex, race/ethnicity, or region of the country.

The VACO index demonstrates that while age is a major driver of COVID-19 mortality, comorbid disease burden quantified by Charlson Comorbidity Index further stratifies risk. The Charlson Comorbidity Index has several strengths in its use as a summary measure of comorbid disease. First, it has been widely demonstrated to be associated with health outcomes in general [13, 14] and specifically in COVID-19 [15]. Second, it can be assessed using EHR data, administrative data, or direct patient interview [13, 16]. Third, the Charlson Comorbidity Index accounts for differences in severity of disease processes by weighting metastatic cancer, AIDS, severe liver disease and diabetes with complications more heavily than other conditions. Fourth, the Charlson Comorbidity Index offers stable weights for conditions that likely contribute to the comorbid disease burden for an individual, but are less common, e.g., rheumatological conditions. When we evaluated whether separately modeling each of the 17 conditions included in the Charlson Comorbidity Index or the Charlson Index score was a better predictor, the AUC in models using the individual conditions were improved by 0.01. However, the weighting assigned to the individual conditions varied more across cohorts than did the weighting assigned the Charlson Index score (data not otherwise shown). Thus a model fit to individual conditions would be less likely to generalize well to new data.

Given the ready accessibility of the VACO Index on MDCalc, we anticipate several immediate applications. The VACO Index may be important for those 60-74 years of age who may both be at substantial risk and often remain employed. In fact, 39% of those age 60-74 in the US are employed [17], thus accurate personalized risk estimation can better inform personal and system level decisions regarding return to work or other group settings. Another important question is how to manage the majority of those undergoing testing in drive up settings. A follow-up phone call to notify the individual that they are positive currently entails a few questions about fever and shortness of breath. If neither are present the patient is advised to quarantine for 2 weeks. However, some of these individuals may be at high risk for serious illness or mortality, justifying a more complete clinical evaluation incorporating vital signs, diagnostic imaging, and laboratory tests. As vaccines and therapeutic options may be in limited supply, the VACO Index could help health systems and government agencies develop a data-driven approach to allocating resources to those at greatest risk of mortality who are most likely to benefit.

Minority populations experienced a greater burden of infection in the beginning of the pandemic [18], leading to suggestions that overall population risk may seem a more equitable criteria for vaccine prioritization. However, modeling population risk of mortality from COVID-19 encompasses two distinct components: probability of infection (infection detection dependent on access to testing and probability of a positive test), and probability of dying once infected. The associations between risk factors such as age, race, and ethnicity are very different for being tested, testing positive, and mortality [18]. The geographic distribution of the pandemic has shifted rapidly over time and testing positive is a volatile outcome that has fluctuated with local disease prevalence. Early in the pandemic in the Spring of 2020 those living in US urban centers on the East and West coasts were at greatest risk of infection. Over time, the pandemic surged in the Midwest and South and extended to rural settings. The second wave of the pandemic in Autumn and Winter of 2020 has largely generalized, with increased infections among Whites and younger individuals. We have demonstrated that risk of mortality after infection has been more stable over time than who is tested and who tests positive. Over the next 6-12 months, nearly everyone not immunized or previously infected is at risk of infection. Thus, we argue that the best strategy is to prioritize those at greatest risk of mortality if infected.

Given the growing debate over the roles of BMI and race/ethnicity in COVID-19 outcomes, we carefully investigated their associations with mortality in our data. Including BMI in the index did not improve the discrimination of the Index as a whole in any of the three data sets in our study. A prospective cohort study of 1,150 COVID-19 inpatients [19], a retrospective cohort of 1,687 inpatients in New York City [20], and a retrospective registry of 331 COVID-19 patients in Italy [21] also did not find an independent association between obesity and mortality. However, in an analysis of 6,916 SARS-CoV-2 positive inpatients and outpatients, Tartof *et al*. did find higher relative risk of death among obese patients [22]. The study authors comment that this increased risk is not uniform across different populations, as the association between high BMI was strong in younger adults ≤ 60 and males. Results from the American Health Association COVID-19 Cardiovascular Disease Registry [23] also had similar findings where they found obese patients to be at higher risk of COVID-19 mortality, notably among adults <50 years and weakly in >70 years. The difference with our findings may be due to the older patient population studied in our samples (VA and Medicare), as age is a major driver in mortality risk, and the number of comorbidities that can account for risk increases with age.

Adding race/ethnicity to the VACO Index did not improve the accuracy of the predictions. Further, associations with race/ethnicity varied among the cohorts, with Black race and Hispanic ethnicity associated with reduced risk of mortality in VA and YNHH data and increased risk among Medicare patients. Since race and ethnicity are largely proxies for socioeconomic disparities [24], it is not surprising that their association with mortality among those testing positive for SARS-CoV-2 is variable. National VA data frequently demonstrates fewer disparities by race in health outcomes than outside VA, likely due to reduced economic barriers to care and national efforts to ensure quality of care [25, 26]. YNHH is a tertiary care academic hospital that draws from Southern Connecticut and Western Rhode Island. In contrast, national Medicare data encompasses widely diverse medical facilities, and minority patients are more likely to be cared for in lower quality facilities [27, 28]. While we considered offering an adjustment for race/ethnicity to be applied outside VA, we are concerned that adding race/ethnicity would introduce substantial noise in the risk estimation due to variation in association across facilities and would confuse biology and socio economics [24, 29]. Importantly, the VACO Index demonstrated consistent or better accuracy in racial and ethnic minority patients as in White patients.

The discrimination of the VACO Index quantified by AUC in Medicare patients was lower than in VA or YNHH patients overall. This discrepancy is related to the reciprocal relationship between the proportion of outcomes events in a population and AUC − all else equal, the AUC will be higher in populations where the outcome is less common [30, 31]. Similarly, when a variable such as age is a major driver of risk, restricting the population tested to a particular age group (e.g., 65 years and over) will result in a constrained AUC [30, 31]. Importantly, the AUC was highly consistent in patients 65 years and over across the three samples and calibration curves in Medicare data demonstrated good agreement between observed and expected mortality rates across diverse races and ethnicities.

Vaccine supply will be limited over the next 6-12 months. We will need a reasonable approach to prioritizing vaccinations in the general population. To minimize lives lost in a generalized epidemic we should vaccinate those at greatest risk of mortality first. The VACO Index shows that for individuals under the age of 85 years, burden of comorbid disease substantially differentiates risk of mortality. For example, a 55-year-old male with a Charlson Comorbidity Index of 10 and peripheral vascular disease has the same mortality risk as an average 65-year-old.

There are limitations to our study. While we were able to evaluate the index in a national sample of over 427,000 people over the age of 65, our ability to validate in substantially younger patients remains somewhat limited. None of the Medicare sample, only 25% of the VA sample, and 20% of the YNHH samples were under 50 years of age. However, the most pressing questions concerning vaccine allocation are focused on individuals over 50 years of age. Also, our calibration is based on mortality events occurring before August 2020 when mostly symptomatic patients were being tested. The VACO Index may over-estimate risk of mortality among asymptomatic patients and this will require further study. Nevertheless, the VACO Index represents the most widely validated risk index for short term mortality from COVID-19. This standardized and validated index can supplement clinical judgement to help inform patient management and health policy.

## Data Availability

The United States Department of Veterans Affairs (VA) places legal restrictions on access to veterans health care data, which includes both identifying data and sensitive patient information. The analytic data sets used for this study are not permitted to leave the VA firewall without a Data Use Agreement. This limitation is consistent with other studies based on VA data. VA data are made freely available to researchers behind the VA firewall with an approved VA study protocol. For more information please visit https://www.virec.research.va.gov or contact the VA Information Resource Center (VIReC) at.

## Supporting information

**S1 Checklist. TRIPOD checklist**.

**S1 Table. Charlson Comorbidity Index Determination from ICD-10 Diagnosis Codes**.

**S1 File. VACO Index Estimation of COVID-19 30-Day Predicted Mortality**.

