## Supplementary materials for "Accuracy of the Veterans Health Administration COVID-19 (VACO) Index for predicting short-term mortality among 1,307 Yale New Haven Hospital inpatients and 427,224 Medicare patients"

### S1. Checklist. TRIPOD Checklist

| Section/Topic | | Checklist Item | Page |
|---|---|---|---|
| **Title and abstract** | | | |
| Title | 1 | Identify the study as developing and/or validating a multivariable prediction model, the target population, and the outcome to be predicted. | Title Page |
| Abstract | 2 | Provide a summary of objectives, study design, setting, participants, sample size, predictors, outcome, statistical analysis, results, and conclusions. | Abstract |
| **Introduction** | | | |
| Background and objectives | 3a | Explain the medical context (including whether diagnostic or prognostic) and rationale for developing or validating the multivariable prediction model, including references to existing models. | Introduction, paragraphs 1-3 |
| | 3b | Specify the objectives, including whether the study describes the development or validation of the model or both. | Introduction, paragraph 4 |
| **Methods** | | | |
| Source of data | 4a | Describe the study design or source of data (e.g., randomized trial, cohort, or registry data), separately for the development and validation data sets, if applicable. | Methods, data source and participants |
| | 4b | Specify the key study dates, including start of accrual; end of accrual; and, if applicable, end of follow-up. | Methods, data source and participants |
| Participants | 5a | Specify key elements of the study setting (e.g., primary care, secondary care, general population) including number and location of centres. | Methods, data source and participants |
| | 5b | Describe eligibility criteria for participants. | Methods, data source and participants |
| | 5c | Give details of treatments received, if relevant. | N/A |
| Outcome | 6a | Clearly define the outcome that is predicted by the prediction model, including how and when assessed. | Methods, data source and participants |
| | 6b | Report any actions to blind assessment of the outcome to be predicted. | Methods, data source and participants |
| Predictors | 7a | Clearly define all predictors used in developing or validating the multivariable prediction model, including how and when they were measured. | Methods, VACO Index components |
| | 7b | Report any actions to blind assessment of predictors for the outcome and other predictors. | Methods, statistical analysis |
| Sample size | 8 | Explain how the study size was arrived at. | Methods, data source and participants |
| Missing data | 9 | Describe how missing data were handled (e.g., complete-case analysis, single imputation, multiple imputation) with details of any imputation method. | Methods, statistical analysis |
| Statistical analysis methods | 10c | For validation, describe how the predictions were calculated. | Methods, statistical analysis |
| | 10d | Specify all measures used to assess model performance and, if relevant, to compare multiple models. | Methods, statistical analysis |
| | 10e | Describe any model updating (e.g., recalibration) arising from the validation, if done. | Methods, statistical analysis |
| Risk groups | 11 | Provide details on how risk groups were created, if done. | Methods, statistical analysis |
| Development vs. validation | 12 | For validation, identify any differences from the development data in setting, eligibility criteria, outcome, and predictors. | Methods, data source and participants |
| **Results** | | | |
| Participants | 13a | Describe the flow of participants through the study, including the number of participants with and without the outcome and, if applicable, a summary of the follow-up time. A diagram may be helpful. | Results, participants |

| | | | |
|---|---|---|---|
| | 13b | Describe the characteristics of the participants (basic demographics, clinical features, available predictors), including the number of participants with missing data for predictors and outcome. | Results, participants |
| | 13c | For validation, show a comparison with the development data of the distribution of important variables (demographics, predictors and outcome). | Results, participants |
| Model performance | 16 | Report performance measures (with CIs) for the prediction model. | Results, risk factors |
| Model-updating | 17 | If done, report the results from any model updating (i.e., model specification, model performance). | Results, VACO Index supplemented |
| **Discussion** | | | |
| Limitations | 18 | Discuss any limitations of the study (such as nonrepresentative sample, few events per predictor, missing data). | Discussion, limitations |
| Interpretation | 19a | For validation, discuss the results with reference to performance in the development data, and any other validation data. | Discussion, paragraphs 1-2 |
| | 19b | Give an overall interpretation of the results, considering objectives, limitations, results from similar studies, and other relevant evidence. | Discussion, paragraphs 1-5 |
| Implications | 20 | Discuss the potential clinical use of the model and implications for future research. | Discussion, paragraph 3 |
| **Other information** | | | |
| Supplementary information | 21 | Provide information about the availability of supplementary resources, such as study protocol, Web calculator, and data sets. | Discussion, paragraph 3 |
| Funding | 22 | Give the source of funding and the role of the funders for the present study. | Funding |

### S1. Table: Charlson Comorbidity Index Determination from ICD-10 Diagnosis Codes

| Comorbidity | ICD-10 Diagnosis Codes | Weight |
|---|---|---|
| AIDS (acquired immunodeficiency syndrome) | B20.x–B22.x, B24.x | 6 |
| Cancer | C00.x–C26.x, C30.x–C34.x, C37.x–C41.x, C43.x, C45.x–C58.x, C60.x–C76.x, C81.x–C85.x, C88.x, C90.x–C97.x | 2 |
| Cancer, metastatic | C77.x–C80.x | 6 |
| Cerebrovascular disease | G45.x, G46.x, H34.0, I60.x–I69.x | 1 |
| Chronic pulmonary disease | I27.8, I27.9, J40.x–J47.x, J60.x–J67.x, J68.4, J70.1, J70.3 | 1 |
| Congestive heart failure (CHF) | I09.9, I11.0, I13.0, I13.2, I25.5, I42.0, I42.5–I42.9, I43.x, I50.x, P29.0 | 1 |
| Dementia | F00.x–F03.x, F05.1, G30.x, G31.1 | 1 |
| Diabetes | E10.0, E10.1, E10.6, E10.8, E10.9, E11.0, E11.1, E11.6, E11.8, E11.9, E12.0, E12.1, E12.6, E12.8, E12.9, E13.0, E13.1, E13.6, E13.8, E13.9, E14.0, E14.1, E14.6, E14.8, E14.9 | 1 |
| Diabetes with complications | E10.2–E10.5, E10.7, E11.2–E11.5, E11.7, E12.2–E12.5, E12.7, E13.2–E13.5, E13.7, E14.2–E14.5, E14.7 | 2 |
| Liver disease, mild | B18.x, K70.0–K70.3, K70.9, K71.3–K71.5, K71.7, K73.x, K74.x, K76.0, K76.2–K76.4, K76.8, K76.9, Z94.4 | 1 |
| Liver disease, moderate or severe | I85.0, I85.9, I86.4, I98.2, K70.4, K71.1, K72.1, K72.9, K76.5, K76.6, K76.7 | 3 |
| Myocardial infarction (MI) | I21.x, I22.x, I25.2, I09.9, I11.0, I13.0, I13.2, I25.5, I42.0, I42.5–I42.9, I43.x, I50.x, P29.0 | 1 |
| Peptic ulcer disease (PUD) | K25.x–K28.x | 1 |
| Peripheral vascular disease (PVD) | I70.x, I71.x, I73.1, I73.8, I73.9, I77.1, I79.0, I79.2, K55.1, K55.8, K55.9, Z95.8, Z95.9 | 1 |
| Plegia | G04.1, G11.4, G80.1, G80.2, G81.x, G82.x, G83.0–G83.4, G83.9 | 2 |
| Renal disease | I12.0, I13.1, N03.2–N03.7, N05.2–N05.7, N18.x, N19.x, N25.0, Z49.0–Z49.2, Z94.0, Z99.2 | 2 |
| Rheumatic disease | M05.x, M06.x, M31.5, M32.x–M34.x, M35.1, M35.3, M36.0 | 1 |

Index calculated by summing comorbidity weights. Three comorbidity pairs are mutually exclusive: Cancer or Cancer, metastatic; Diabetes or Diabetes with complications; and Liver disease, mild or Liver disease, moderate or severe – only use the higher weight, not both.

Adapted from Quan *et al*., Coding algorithms for defining comorbidities in ICD-9 and ICD-10 administrative data. *Med Care* 2005; 43(11): 1130-1139.

### S1. File: VACO Index Calculation of Predicted COVID-19 30-day Mortality

| Variable | Coefficient | Z | P-value | OR | 95% CI |
|---|---|---|---|---|---|
| Age, years | | | | | |
| 20-49 | -2.228678713 | -2.83 | 0.005 | 0.11 | 0.02 - 0.50 |
| 50-54 | 0.0 | | - | 1.00 | - |
| 55-59 | 0.400599289 | 0.97 | 0.334 | 1.49 | 0.66 - 3.36 |
| 60-64 | 0.941322019 | 2.50 | 0.013 | 2.56 | 1.22 - 5.37 |
| 65-69 | 1.295007128 | 3.49 | <0.001 | 3.65 | 1.77 - 7.55 |
| 70-74 | 1.629533438 | 4.55 | <0.001 | 5.10 | 2.53 - 10.3 |
| 75-79 | 1.763345763 | 4.72 | <0.001 | 5.83 | 2.81 - 12.12 |
| 80-89 | 1.927443543 | 4.96 | <0.001 | 6.87 | 3.21 - 14.72 |
| ≥90 | 2.018752269 | 4.39 | <0.001 | 7.53 | 3.06 - 18.54 |
| Sex | | | | | |
| Female | 0.0 | - | - | 1.00 | - |
| Male | 0.322291449 | 0.88 | 0.377 | 1.38 | 0.68 - 2.82 |
| CCI and Age Interaction Term | | | | | |
| Age <85 | | | | | |
| CCI | | | | | |
| 0 | 0.0 | - | - | - | - |
| 1-3 | 0.612122574 | 2.76 | 0.006 | 1.84 | 1.19 - 2.85 |
| 4-5 | 0.825072847 | 3.36 | 0.001 | 2.28 | 1.41 - 3.69 |
| 6-9 | 0.956099733 | 3.84 | <0.001 | 2.60 | 1.60 - 4.24 |
| ≥10 | 1.395164653 | 4.25 | <0.001 | 4.04 | 2.12 - 7.68 |
| Age 85+, any CCI | 1.529325519 | 4.79 | <0.001 | 4.62 | 2.47 - 8.63 |
| MI or PVD | | | | | |
| No | 0.0 | - | - | 1.00 | - |
| Yes | 0.267265312 | 2.18 | 0.029 | 1.31 | 1.03 - 1.66 |
| Constant | -4.216058062 | -8.41 | <0.001 | 0.01 | 0.01 - 0.04 |

Abbreviations: OR = odds ratio; CI = confidence interval; CCI =Charlson comorbidity index; MI = myocardial infarction; PVD = peripheral vascular disease

Calculation of predicted mortality risk:

$\text{coefficient}_{sum} = \text{Age}_{coefficient} + \text{Sex}_{coeffient} + \text{CCI\_Age}_{coefficient} + \text{MI\_PVD}_{coefficient} + \text{Constant}_{coefficient}$

$\text{OR}_{calc} = \exp(\text{coefficient}_{sum})$

$\text{risk}_{pred} = \text{OR}_{calc} / (1 + \text{OR}_{calc})$

Example: 77 year old male with CCI = 5, but no history of MI or PVD

$\text{coefficient}_{sum} = 1.763345763 + 0.322291449 + 0.825072847 + 0.0 + (-4.216058062) = -1.305348004$

$\text{OR}_{calc} = \exp(-1.305348004) = 0.\ 271078182$

$\text{risk}_{pred} = 0.271078182 / (1 + 0.271078182) = 0.213266333 = 21\%$
